# Investigating Racial Disparities in Drug Prescriptions for Patients with Endometriosis

**DOI:** 10.1101/2023.10.02.23296435

**Authors:** Aparajita Kashyap, Maryam Aziz, Tony Y. Sun, Sharon Lipsky-Gorman, Jessica Opoku-Anane, Noémie Elhadad

**Affiliations:** Columbia University Irving Medical Center, Department of Biomedical Informatics (New York, NY); Columbia University, Department of Computer Science (New York, NY); Columbia University Irving Medical Center, Department of Obstetrics and Gynecology (New York, NY)

**Keywords:** Health disparities, Reproductive healthcare, observational study, biomedical informatics, prescription patterns, pain medication, chronic disease, temporal analysis

## Abstract

**Background:** Endometriosis is a chronic disease with a long time to diagnosis and several known comorbidities that requires a range of treatments including of pain management and hormone-based medications. Racial disparities specific to endometriosis treatments are unknown.

**Objective:** We aim to investigate differences in patterns of drug prescriptions specific to endometriosis management in Black and White patients prior to diagnosis and after diagnosis of endometriosis and compare these differences to racial disparities established in the general population.

**Study Design:** We conduct a retrospective cohort study using observational health data from the IBM MarketScan® Multi-state Medicaid dataset. We identify a cohort of endometriosis patients consisting of women between the ages of 15 and 49 with an endometriosis-related surgical procedure and a diagnosis code for endometriosis within 30 days of this procedure. Cohort is further restricted to patients with at least 3 years of continuous observation prior to diagnosis.

We identify a non-endometriosis cohort of women between the ages of 15 and 49 with no endometriosis diagnosis and at least 1 year of continuous observation. We compare prevalence of prescriptions across selected drug classes for Black vs. White endometriosis patients. We further examine prevalence differences in the non-endometriosis cohort and prevalence differences pre- and post-diagnosis in the endometriosis cohort.

**Results:** The endometriosis cohort comprised 16,372 endometriosis patients (23.3% Black, 66.0% White). Of the 28 drug classes examined, 17 were prescribed significantly less in Black patients compared to 21 in non-endometriosis cohort (n=3,663,904), and 4 were prescribed significantly more in Black patients compared to 6 in the non-endometriosis cohort. Of the 17 drugs prescribed more often in White patients, 16 have larger disparities pre-diagnosis than post-diagnosis.

**Conclusions:** Our analysis identified significant differences in medication prescriptions between White and Black patients with endometriosis, notably in hormonal treatments, pain management, and treatments for common endometriosis co-morbidities. Racial disparities in drug prescriptions are well established in healthcare, and better understanding these disparities in the specific context of chronic reproductive conditions and chronic pain is important for increasing equity in drug prescription practices.

## Introduction

Endometriosis is a chronic, inflammatory disease characterized by growth of endometrial-like tissue outside the uterus. Identified symptoms of endometriosis vary widely and include dysmenorrhea, fatigue, non-menstrual abdominopelvic pain, and heavy menstrual bleeding^1,2^. While the precise prevalence of endometriosis is unknown, it affects an estimated 6-10% of women of reproductive age^2–4^. Current medical interventions for endometriosis depend on patient prognosis and include analgesics, combined hormonal contraceptives, progestogens, gonadotropin-releasing hormone (GnRH) agonists, GnRH antagonists, aromatase inhibitors, and surgical treatments^5^.

Racial health disparities in the United States are pervasive, and research shows that Black and Hispanic patients are less likely to receive a diagnosis of endometriosis compared to White patients^6^, as well as less likely to receive surgical treatment for endometriosis^7^. Racial bias is observed beyond endometriosis diagnosis, such as in pain management; Black and Hispanic patients are less likely to be treated for pain and other symptoms that impact quality of life^8–11^.

In this work we characterize the drug prescription patterns for endometriosis patients and potential differences in prevalence across Black and White patients. Because endometriosis is a condition with delays in diagnosis, patients with suspected endometriosis are often prescribed first-line treatments prior to their diagnosis. We thus characterize treatment patterns in three ways: prior to diagnosis, after diagnosis, and overall. Furthermore, to account for multiple co-morbidities of endometriosis, we extend our analysis to all medication classes that are significantly more prescribed in our endometriosis cohort than in a non-endometriosis comparison cohort. To further contextualize this analysis, we compare treatment patterns of endometriosis patients with those without endometriosis and measure differences in treatment patterns across White and Black patients in a non-endometriosis cohort, namely all women in reproductive age without any diagnosis of endometriosis.

## Materials and Methods

The analysis and use of the de-identified dataset presented in this work were carried out under Research Protocol AAAO7805 approved by the Columbia University IRB.

### Dataset

In this retrospective cohort study, we focus our analysis to patients with Medicaid, the U.S. government program that provides health insurance for people with limited income. The data comes from the IBM MarketScan® Multi-state Medicaid dataset, which draws Medicaid data from several states^12^. The Medicaid dataset contains de-identified longitudinal records of patients and includes inpatient and outpatient services, diagnostic history, and drug prescriptions for over 48 million enrollees^12^. We leverage the data under the OMOP Common Data Model (CDM) format, which follows standardized conventions for drugs, therapies, and other medical vocabularies^13^. The database has been used for a variety of observational health studies due to its flexibility and robustness^14–16^.

### Cohort Identification

We select patients with endometriosis using a validated cohort definition^17^. The cohort definition includes women ages 15 to 49 years who have an endometriosis-related surgical procedure (e.g., laparoscopic surgery) as well as a diagnosis of endometriosis 30 days before or after this procedure. The phenotype definition was validated through manual chart review of 1,400 endometriosis patients yielding 70% sensitivity, 93% specificity, 85% positive predictive value. Age is calculated at time of diagnosis. Only patients with endometriosis with at least three years of continuous observation prior to diagnosis are considered for the endometriosis cohort.

The non-endometriosis cohort comprises of any woman aged 15-49 with no diagnostic code for endometriosis at any point in their medical history and at least one year of continuous observation. Age is calculated using the most recent visit date. In the claims database, it is unknown for individual patients whether the gender marker represents sex, gender, or both.

### Identification of medications

The Anatomical Therapeutic Chemical (ATC) classification system separates drugs in a standardized manner; we create drug classes using ATC level 3 classifications because they are associated with specific pharmacological functions without specifying chemical structures^18,19^. We use the OMOP CDM which explicitly maps medications to their ATC level 3 classification. The ATC classifications create a “bridge” between the clinical guidelines for endometriosis and large-scale observational health data.

We narrow the set of drug classes by identifying drug classes that are prescribed significantly more often in the endometriosis cohort than the non-endometriosis one. For each drug class, the proportion of patients in each cohort with at least one prescription belonging to that drug class is calculated. The statistical significance of the difference between these relative prevalence measurements is calculated using a two-sided Z-test with a 0.01 significance level. This procedure identified 28 drug classes that are significantly more prevalent in the endometriosis cohort than the non-endometriosis cohort (see **Supplemental Table 1**); we then measure prescription prevalence for each of these drug classes.

To understand the clinical relevance of these 28 drug classes, we checked them against the drugs listed in the 2022 ESHRE guidelines for treating endometriosis^5^. The only recommended treatments that are not included in our analysis are GnRH antagonists and aromatase inhibitors. GnRH antagonists were not prescribed in the Medicaid dataset, and aromatase inhibitors were excluded because the corresponding ATC 3^rd^ level drug class (“hormone antagonists and related agents”) was not prescribed significantly more in patients with endometriosis than in the non-endometriosis cohort. Several other drug classes more prevalently prescribed in the endometriosis population are associated with known comorbidities or symptoms of endometriosis including gastrointestinal distress, depression, and anxiety^23,24^.

### Temporal Analysis

Endometriosis is known to have extended delays between symptom onset and diagnosis^20,21^. We therefore aim to better understand the patterns of drug prescription prior to diagnosis (which occurs via laparoscopic surgery) and whether they differ from prescription practices after diagnosis. Prescription practices from the pre-diagnosis period can be thought of as prescription practices for undiagnosed endometriosis patients, which is an important area of study given that Black patients are less likely to be diagnosed with endometriosis compared to White patients^6^.

We create two temporal subgroups of drug prescriptions in the endometriosis cohort (see **Figure 1**). Subgroup 1 consists of all prescriptions occurred prior to surgical diagnosis. Subgroup 2 consists of all prescriptions post-diagnosis. Note that neither subgroup 1 nor subgroup 2 contain prescriptions given on the day of diagnosis; these prescriptions are only counted in the “overall” prescriptions.

**Figure 1:**
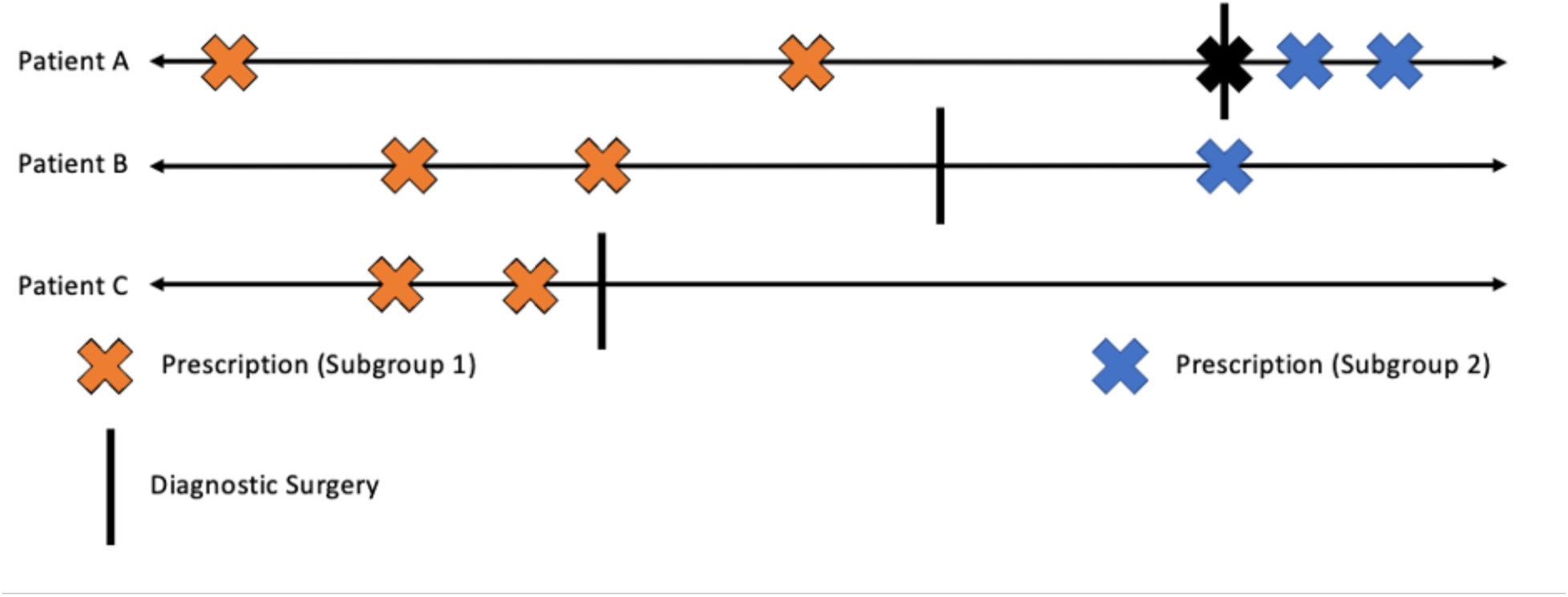
Temporal subgroups for patient prescriptions. Three example longitudinal records are shown, with prescriptions marked as “x” and a laparoscopic diagnosis of endometriosis marked with a vertical line. As shown in the figure, prescriptions ordered prior to diagnosis fall into subgroup 1 (pre-diagnosis, orange) and prescriptions ordered after diagnosis fall into subgroup 2 (post-diagnosis, blue). Prescriptions from the day of diagnosis are not counted in either subgroup.

### Statistical Methods

For each drug class of interest, the relative prevalence of prescriptions within that drug class was calculated for Black and White endometriosis patients. A person is counted as part of the drug-positive group if they received at least one prescription that fell into the given drug class, and relative prevalence is calculated based on the total number of Black (n = 3,814) and White (n = 10,805) patients in the endometriosis cohort. The difference in prescription prevalence is calculated as 100% ∗ (*p*_*white*_ − *p*_*black*_), where *p*_*white*_ and *p*_*black*_ relative prevalence (see **Table 2**, **Table 3**).

To identify significant differences in the Black and White sub-populations, we conduct a two-sided Z-test from the difference in prevalence and the associated standard error. We apply the Bonferroni correction to account for multiple comparisons; we consider the difference significant when the adjusted p-value < 0.01. This procedure for statistical analysis was repeated for all prescription classes and both temporal subgroups (pre-diagnosis and post-diagnosis). Analysis was carried out using the scipy package in Python^22^.

## Results

### Cohorts

The non-endometriosis cohort comprises 3,663,904 patients (see **Table 1**). The endometriosis cohort (n=16,372) comprised more White (66%) than the non-endometriosis cohort (23%) (of note, 50% of patients in the non-endometriosis cohort have no recorded race). The endometriosis cohort also was slightly older (mean age 32.4 ± 8.0 years) than the non-endometriosis cohort (mean age 28.7 ± 10.3 years). The median length of observational period was 6.08 (IQR: 5.08 to 8.84) years for the endometriosis cohort and 4.00 (IQR: 2.00 to 6.25) years for the non-endometriosis cohort. For the endometriosis cohort, the median length of observation prior to diagnosis was 4.20 (IQR: 3.26 to 5.73) years, and the median length of observation post-diagnosis was 1.68 (IQR: 0.82 to 2.89) years.

**Table 1:**
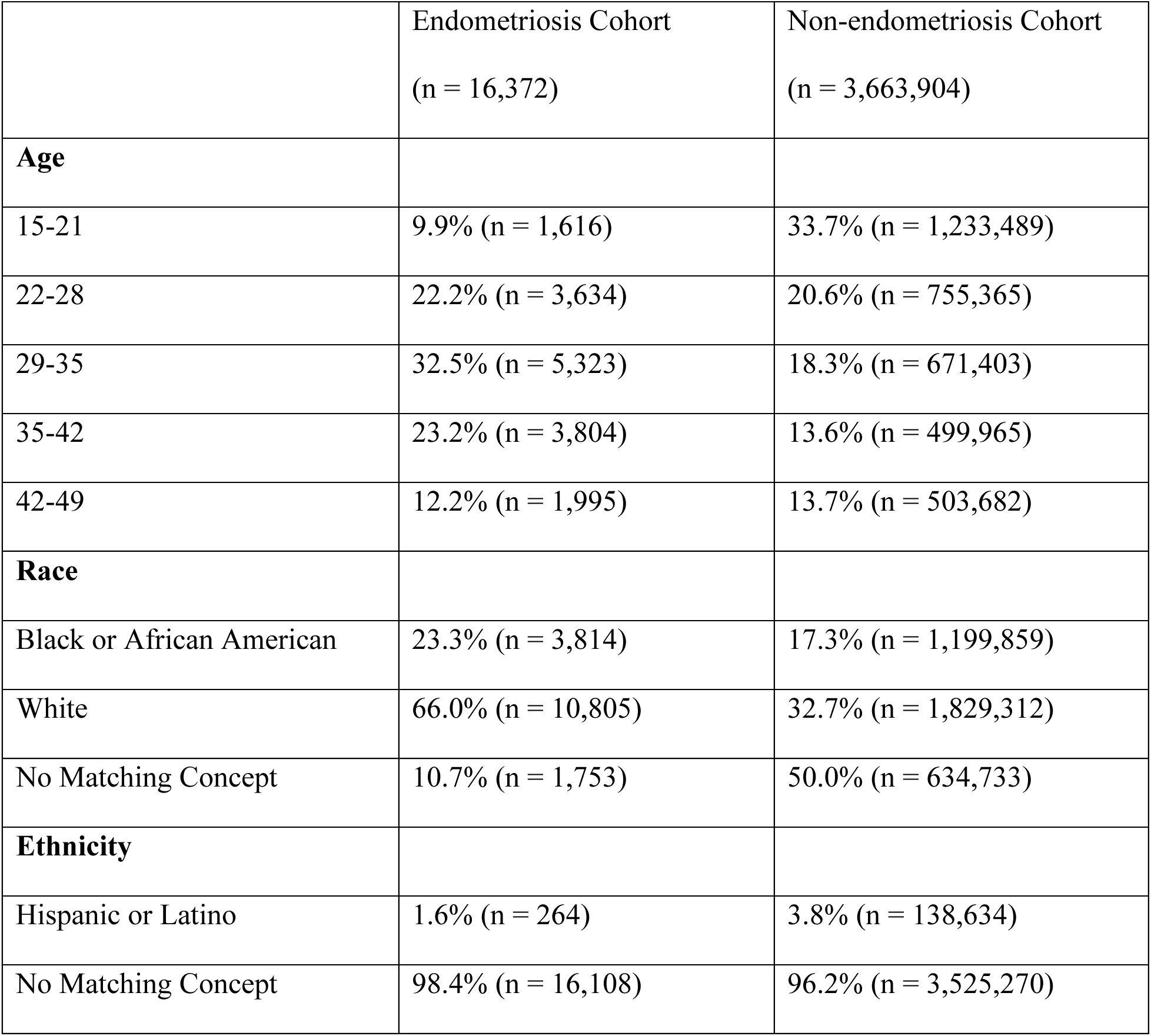
Descriptive statistics of the endometriosis and non-endometriosis cohorts. This table contains information about the age, race and ethnicity of patients in the endometrsiosi and non-endometriosis cohorts. Age is calculated as the date of cohort entry (date of diagnosis for endometriosis patients, date of most recent visit for non-endometriosis patients).

|  | Endometriosis Cohort<br>(n = 16,372) | Non-endometriosis Cohort<br>(n = 3,663,904) |
| --- | --- | --- |
| **Age** |  |  |
| 15-21 | 9.9% (n = 1,616) | 33.7% (n = 1,233,489) |
| 22-28 | 22.2% (n = 3,634) | 20.6% (n = 755,365) |
| 29-35 | 32.5% (n = 5,323) | 18.3% (n = 671,403) |
| 35-42 | 23.2% (n = 3,804) | 13.6% (n = 499,965) |
| 42-49 | 12.2% (n = 1,995) | 13.7% (n = 503,682) |
| **Race** |  |  |
| Black or African American | 23.3% (n = 3,814) | 17.3% (n = 1,199,859) |
| White | 66.0% (n = 10,805) | 32.7% (n = 1,829,312) |
| No Matching Concept | 10.7% (n = 1,753) | 50.0% (n = 634,733) |
| **Ethnicity** |  |  |
| Hispanic or Latino | 1.6% (n = 264) | 3.8% (n = 138,634) |
| No Matching Concept | 98.4% (n = 16,108) | 96.2% (n = 3,525,270) |

### Differences in overall prevalence of drug prescriptions for Black and White patients

Of the 28 drug classes of interest, 17 (61%) are prescribed significantly more often for White endometriosis patients compared to Black endometriosis patients (see **Figure 2**). In the non-endometriosis cohort, 21 (75%) are prescribed significantly more for White patients than for Black patients and 6 (21%) are prescribed significantly more for Black patients.

**Figure 2:**
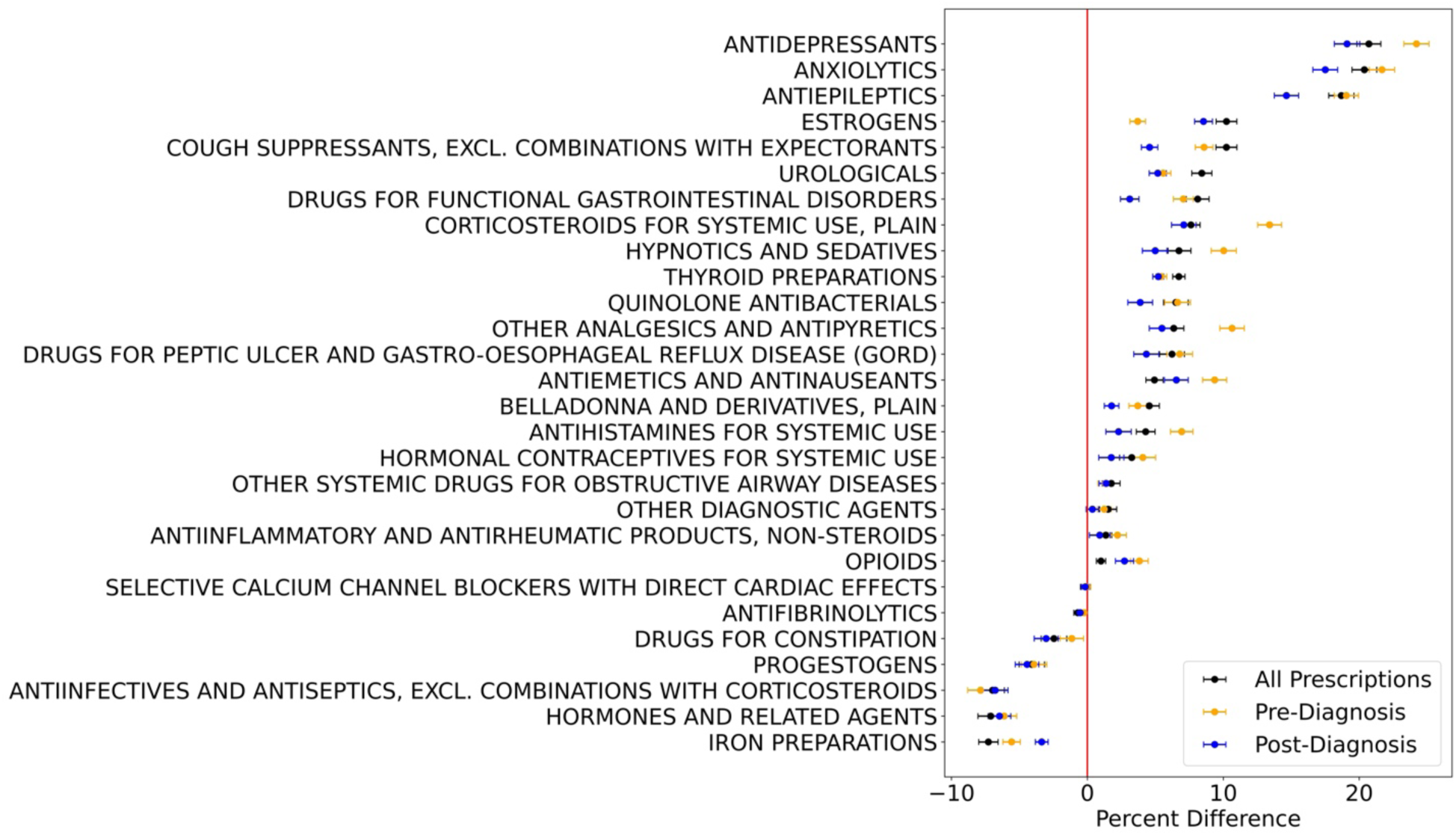
Difference in drug class prescription prevalence for White and Black patients. This graph shows the percent difference in drug class prescription prevalence between White and Black patients. Error bars indicate standard error, and a difference greater than 0 indicates that White patients are prescribed a drug from that class with higher prevalence than Black patients. The percent difference is shown across all visits, visits prior to diagnosis (subgroup 1), and visits after diagnosis (subgroup 2).

The largest racial differences in endometriosis patients occur for antidepressants (prescription prevalence higher by 20.7% in White patients than Black patients, 95% CI [19.8%, 21.6%]), anxiolytics (20.4%, 95% CI [19.5%, 21.3%]), antiepileptics (18.7%, 95% CI [17.8%, 19.6%]), and estrogens (10.2%, 95% CI [9.5%, 11.0%]). Additionally, 4 drug classes are prescribed significantly more often in Black patients than White patients, with the largest differences occurring for iron preparations (7.3%, 95% CI [6.6%, 8.0%]), hormones and related agents (7.1%, 95% CI [6.2%, 8.1%]), and antiinfectives/antiseptics (7.0%, 95% CI [6.1%, 7.9%]). Prescription prevalence differences are more pronounced in the endometriosis cohort than the non-endometriosis cohort for anxiolytics, antiepileptics, estrogens, iron preparations, and antiinfectives/antiseptics but are less pronounced in the endometriosis cohort for antidepressants and hormones and related agents (see **Table 2**).

**Table 2:** Overall prescription prevalence in the endometriosis and the non-endometriosis cohort. This table shows the differences in prescription prevalence (mean prevalence difference) for White and Black patients in the endometriosis and non-endometriosis cohorts. The standard error and adjusted p-value associated with the difference in prevalence is also reported. Significant p-values (p < 0.01) are emphasized in bold.

| Drug Class | Endometriosis Cohort |  |  | Non-endometriosis Cohort |  |  |
| --- | --- | --- | --- | --- | --- | --- |
|  | Mean Prevalence Difference (%) | Standard Error (%) | Adjusted p-value | Mean Prevalence Difference (%) | Standard Error (%) | Adjusted p-value |
| Antidepressants | 20.7092674 | 0.8779937 | **7.33E-122** | 22.9035692 | 0.05304433 | **0** |
| Anxiolytics | 20.3798907 | 0.9113823 | **1.31E-109** | 15.6527752 | 0.05042332 | **0** |
| Antiepileptics | 18.6863978 | 0.92753865 | **4.07E-89** | 12.9321962 | 0.04436682 | **0** |
| Estrogens | 10.2351817 | 0.76278335 | **6.62E-40** | 1.8295592 | 0.02425678 | **0** |
| Cough suppressants, excl. Combinations with expectorants | 10.2306415 | 0.7721378 | **6.34E-39** | 5.98557177 | 0.03181319 | **0** |
| Urologicals | 8.41948621 | 0.73720344 | **4.61E-29** | 2.94880655 | 0.02595341 | **0** |
| Drugs for functional gastrointestinal disorders | 8.11512033 | 0.83853302 | **5.25E-21** | 2.98066657 | 0.02871509 | **0** |
| Corticosteroids for systemic use, plain | 7.61934586 | 0.67395744 | **1.73E-28** | 12.0253099 | 0.05781293 | **0** |
| Hypnotics and sedatives | 6.73798789 | 0.88230912 | **3.12E-13** | 7.11677719 | 0.04465059 | **0** |
| Thyroid preparations | 6.72891005 | 0.44941487 | **1.55E-49** | 4.43773974 | 0.02185524 | **0** |
| Quinolone<br>antibacterials | 6.51072657 | 0.90775091 | **1.03E-11** | 6.41267606 | 0.04888034 | **0** |
| Other analgesics and<br>antipyretics | 6.35556622 | 0.7432395 | **1.71E-16** | 7.47203815 | 0.05709184 | **0** |
| Drugs for peptic ulcer<br>and gastro-oesophageal<br>reflux disease (GORD) | 6.22370589 | 0.91284682 | **1.29E-10** | 7.15618371 | 0.05234589 | **0** |
| Antiemetics and<br>antinauseants | 4.93577208 | 0.63104266 | **7.30E-14** | 7.03386181 | 0.05758966 | **0** |
| Belladonna and<br>derivatives, plain | 4.5541342 | 0.74166983 | **1.15E-08** | 2.25201094 | 0.02541702 | **0** |
| Antihistamines for<br>systemic use | 4.28910075 | 0.69098553 | **7.55E-09** | 3.91808881 | 0.05868259 | **0** |
| Hormonal<br>contraceptives for<br>systemic use | 3.26120892 | 0.88940093 | **0.00343917** | -1.9807205 | 0.05820126 | **1.04E-252** |
| Other systemic drugs<br>for obstructive airway<br>diseases | 1.75543863 | 0.64534074 | 0.0913468 | 1.57064744 | 0.02842173 | **0** |
| Other diagnostic agents | 1.52705867 | 0.62911922 | 0.21296566 | 0.67426432 | 0.0239525 | **3.35E-173** |
| Antiinflammatory and<br>antirheumatic products,<br>non-steroids | 1.35203434 | 0.43286802 | 0.02502562 | 1.17594879 | 0.0579422 | **1.98E-90** |
| Opioids | 1.00063649 | 0.3378741 | 0.04285102 | 4.93323827 | 0.05817166 | **0** |
| Selective calcium<br>channel blockers with<br>direct cardiac effects | -0.1429571 | 0.36793811 | 9.76667522 | 0.14467697 | 0.01170009 | **5.63E-34** |
| Antifibrinolytics | -0.7107767 | 0.28571871 | 0.18001115 | 0.00790219 | 0.00613992 | 2.77322788 |
| Drugs for constipation | -2.4573705 | 0.94071259 | 0.12593019 | -1.0426026 | 0.04477045 | **8.26E-119** |
| Progestogens | -4.0913806 | 0.93452218 | **0.00016765** | -7.8743071 | 0.05176947 | **0** |
| Antiinfectives and<br>antiseptics, excl.<br>Combinations with<br>corticosteroids | -6.9788283 | 0.87299686 | **1.83E-14** | -6.3718204 | 0.0524945 | **0** |
| Hormones and related<br>agents | -7.1121568 | 0.9398425 | **5.33E-13** | -8.9228181 | 0.04865653 | **0** |
| Iron preparations | -7.2843687 | 0.71144019 | **1.86E-23** | -4.4471093 | 0.03375195 | **0** |

Out of the drug classes related to established endometriosis treatment options, three (opioids, other analgesics and antipyretics, and hormonal contraceptives for systemic use) are significantly more prevalent amongst White endometriosis patients; the remaining two (hormones and related agents, progestogens) are significantly more prevalent amongst Black endometriosis patients (see **Table 2**). These differences are generally consistent with the non-endometriosis cohort, where analgesics are more commonly prescribed for White women and hormones and related agents are more commonly prescribed in Black women. One major difference between the endometriosis and non-endometriosis cohorts, however, is in hormonal contraceptives for systemic use, which are prescribed significantly more in White women with endometriosis (3.3%) and significantly more for Black women without endometriosis (2.0%). Additionally, the difference in prescriptions of estrogens is much higher in the endometriosis cohort (10.23%) than in the non-endometriosis cohort (1.83%).

### Temporal differences in drug prescription differences

When observing drug prescription differences across White and Black endometriosis patients across temporal subgroups, there is a pattern of pre-diagnosis differences (temporal subgroup 1) exceeding post-diagnosis differences (temporal subgroup 2), especially for drug classes where White patients are prescribed at a higher rate (see **Figure 2** and **Table 3**). Of the 17 drug classes that fall into this category, 16 (94%) have larger differences prior to diagnosis compared to post-diagnosis. For drugs that are significantly more prevalently prescribed in Black patients, the difference in prevalence decreases from pre- to post-diagnosis for iron preparations and antiinfectives/antiseptics; the difference in prevalence is greater post-diagnosis for hormones and related agents and progestogens.

**Table 3:** Disparities in Prescription Prevalence Pre-Diagnosis and Post-Diagnosis. This table contains all 28 drug classes and the difference in prevalence between White patients and Black patients. The associated standard error and p-value are also reported. A positive mean prevalence difference indicates that White patients were prescribed a drug within that class more often. Significant p-values (p < 0.01) are emphasized in bold.

| Drug Class | Pre-Diagnosis |  |  | Post-Diagnosis |  |  |
| --- | --- | --- | --- | --- | --- | --- |
|  | Mean Prevalence Difference (%) | Standard Error (%) | Adjusted p-value | Mean Prevalence Difference (%) | Standard Error (%) | Adjusted p-value |
| Antidepressants | 24.2136409 | 0.91910921 | **8.27E-152** | 19.1033773 | 0.93308442 | **5.21E-92** |
| Anxiolytics | 21.6790021 | 0.92689508 | **7.77E-120** | 17.5023318 | 0.91112685 | **4.31E-81** |
| Antiepileptics | 19.0576737 | 0.8894292 | **1.05E-100** | 14.6465251 | 0.89299566 | **2.61E-59** |
| Corticosteroids for systemic use, plain | 13.4112531 | 0.88354112 | **6.82E-51** | 7.09665138 | 0.90348168 | **5.61E-14** |
| Other analgesics and antipyretics | 10.647111 | 0.89842351 | **2.98E-31** | 5.48276992 | 0.9261711 | **4.51E-08** |
| Hypnotics and sedatives | 10.0331305 | 0.92767679 | **4.08E-26** | 4.99233052 | 0.94689986 | **1.89E-06** |
| Antiemetics and antinauseants | 9.36654584 | 0.88938431 | **8.66E-25** | 6.54327582 | 0.87627634 | **1.15E-12** |
| Cough suppressants, excl. Combinations with expectorants | 8.58926626 | 0.64989361 | **9.88E-39** | 4.5777826 | 0.60477066 | **5.25E-13** |
| Drugs for functional gastrointestinal disorders | 7.06331071 | 0.72801319 | **4.13E-21** | 3.11520408 | 0.68196701 | **6.89E-05** |
| Antihistamines for systemic use | 6.94421682 | 0.82975723 | **8.14E-16** | 2.29503444 | 0.93040888 | 0.19091332 |
| Drugs for peptic ulcer and gastro-oesophageal reflux disease (GORD) | 6.79679748 | 0.94385519 | **8.36E-12** | 4.34071438 | 0.92647094 | **3.92E-05** |
| Quinolone antibacterials | 6.63919703 | 0.94358424 | **2.77E-11** | 3.88632803 | 0.91046271 | **0.00027548** |
| Urologicals | 5.58218552 | 0.56071233 | **3.34E-22** | 5.18144241 | 0.62964183 | **2.64E-15** |
| Thyroid preparations | 5.44076752 | 0.40182738 | **1.27E-40** | 5.21899767 | 0.40262683 | **2.80E-37** |
| Hormonal contraceptives for systemic use | 4.08447171 | 0.93484337 | **0.00017462** | 1.75921784 | 0.92248195 | 0.79120025 |
| Opioids | 3.83223567 | 0.64179058 | **3.30E-08** | 2.73050637 | 0.66811974 | **0.00061217** |
| Belladonna and derivatives, plain | 3.71041927 | 0.64993182 | **1.59E-07** | 1.78282791 | 0.53391252 | 0.01176382 |
| Estrogens | 3.70365484 | 0.5754246 | **1.71E-09** | 8.54471765 | 0.65909065 | **2.73E-37** |
| Antiinflammatory and antirheumatic products, non-steroids | 2.21986375 | 0.65859413 | 0.01050029 | 0.90768334 | 0.76028642 | 3.25540217 |
| Other systemic drugs for obstructive airway diseases | 1.3305698 | 0.54676884 | 0.20934261 | 1.38698549 | 0.52434092 | 0.11430042 |
| Other diagnostic agents | 1.24341988 | 0.51596245 | 0.22339418 | 0.37362264 | 0.45886946 | 5.81723098 |
| Selective calcium channel blockers with direct cardiac effects | -0.0672012 | 0.31416557 | 11.628708 | -0.1512507 | 0.26786714 | 8.01238248 |
| Antifibrinolytics | -0.2432193 | 0.22567947 | 3.9362227 | -0.5235776 | 0.19670476 | 0.10883203 |
| Drugs for constipation | -1.1445405 | 0.87089075 | 2.6428188 | -3.0364077 | 0.8782654 | **0.00763926** |
| Progestogens | -3.9202865 | 0.93975007 | **0.00042341** | -4.4411163 | 0.87031541 | **4.68E-06** |
| Iron preparations | -5.5722235 | 0.64193868 | **5.53E-17** | -3.3586467 | 0.4702192 | **1.28E-11** |
| Hormones and related<br>agents | -6.1206816 | 0.92499823 | **5.13E-10** | -6.4695481 | 0.8338995 | **1.21E-13** |
| Antiinfectives and<br>antiseptics, excl.<br>combinations with<br>corticosteroids | -7.8604257 | 0.9433205 | **1.11E-15** | -6.7822939 | 0.94208523 | **8.48E-12** |

## Comment

### Principal Findings

Our analysis of drug prescription patterns amongst White and Black patients with endometriosis in the Medicaid population shows racial disparities in disease treatment and pain management within endometriosis. Differences in prescription prevalence tend to be larger pre-diagnosis compared to post-diagnosis. We also find that drugs associated with comorbidities of endometriosis (including gastrointestinal distress, irritable bowel syndrome, thyroid disorders, and lower urinary tract symptoms^25–27^) are prescribed significantly less prevalently in Black patients compared to White patients, often with larger disparities in the endometriosis population than the non-endometriosis population; this suggests disparities in holistic endometriosis management.

### Results in the Context of What is Known

Several disparities identified in this dataset are well-documented in patient populations not specific to endometriosis (e.g. prescriptions of antidepressants^28,29^, select anxiolytics^30^, and pain medication^8–11^). Prior studies have also observed racial disparities in endometriosis diagnosis^6^ and surgical treatments^7^; therefore, it becomes important to consider how lack of treatment and lack of diagnosis may collectively impact patients’ healthcare experiences. Additionally, when considering that most prescription disparities are larger pre-diagnosis compared to post-diagnosis, we conceptualize the pre-diagnosis period as one where the patient may require treatment but has no formal diagnosis to explain the multi-system, chronic symptoms they may be facing.

### Clinical Implications

The most notable differences in prescription prevalence occur for antidepressants and anxiolytics, which are generally used to treat depression and anxiety. Depression and anxiety are known comorbidities of endometriosis and are associated with worse endometriosis symptoms^23,24^. Prior studies have found that Black patients experience less rapport-building during visits^31^ and are additionally less likely to communicate their symptoms (especially those related to mental health) to healthcare providers^31,32^; this suggests a gap in physician-patient communication which could be due to lack of cultural competency and/or perceived racism^33^. Perceived racism^34^ includes acts or attitudes that may or may not “objectively” considered racist but do contribute to the (subjective) experience of racial prejudice and is known to contribute to healthcare-related stress^35–37^. Future qualitative work should investigate how endometriosis patients (who typically have more frequent interactions with the healthcare system) experience racism and the impact this has on their healthcare-related stress.

We observe disparities in the prescription of pain medication (both opioid and non-opioid) where Black patients are less likely to receive this treatment than White patients. False racial assumptions and stereotypes such as the idea that Black patients asking for pain medications are “drug-seeking”^38,39^ or that Black patients feel less pain than their White counterparts^8^ may contribute to these disparities. Perceived racism can also impact how Black patients experience pain management: in addition to exacerbating healthcare-related stress, the racism experienced by Black patients when seeking chronic pain treatment may also contribute to hopelessness (negative expectations about one’s present life and future), which can negatively impact pain management^10^. Additionally, misconceptions and lack of effective communication between patients and physicians surrounding medication tolerance, side effects, efficacy, and addiction has been found to disproportionately impact racial and ethnic minorities^37^. Our finding that the disparities in pain medication prescriptions is greater in the pre-diagnosis cohort than the post-diagnosis subgroups suggests that the reasons for lack of prescriptions are exacerbated pre-diagnosis when patients have symptoms of, but are not explicitly diagnosed with, endometriosis compared to post-diagnosis, when their endometriosis has been formally diagnosed.

Finally, when comparing racial prevalence disparities across the endometriosis and non-endometriosis cohorts, there was broad consistency; however, several of the drug classes associated with comorbidities of endometriosis (anxiolytics, urologicals, drugs for functional gastrointestinal disorders, thyroid preparations) had disparities that were larger in the endometriosis than the non-endometriosis cohort. There may be inequitable treatment of Black women specific to endometriosis, especially during their journey to diagnosis (when all these disparities are larger). Notably, the medication class with the highest difference was Estrogens (see **Table 2**). There are potentially multiple indications to drugs under that medication class that may differ in the endometriosis and non-endometriosis cohorts, such as hormone replacement therapy for post-menopausal women vs. fertility treatments for endometriosis patients.

### Research Implications

We identify several classes of drugs where there may be racial disparities in prescriptions specifically for endometriosis treatment; however, this work does not dive into the context of these drug prescription patterns, nor does it investigate these disparities in the context of patient experiences and quality of life. Future work should focus on better understanding the context of drug prescription patterns, especially co-occurring conditions associated with specific prescriptions or self-reported data about pain and how that relates to analgesic prescriptions for White and Black patients. Additionally, qualitative work should be conducted to understand the multifactorial reasons for our findings, to assess the impact of these disparities on patient lives, and to identify methods for mitigating these disparities. Finally, this analysis should be extended to other patient populations to identify disparities in other racial and ethnic minorities, as well as uninsured patients (who may face even more barriers to care than the Medicaid patients in this dataset).

### Strengths and Limitations

One major strength of this study was the use of the Multi-state Medicaid Dataset. Medicaid covers 72.5 million Americans, making it the largest source of healthcare in the United States^40^. Furthermore, in this study we aimed to compare groups of patients with similar access to care (e.g., patients with at least three years of continuous observation prior to endometriosis diagnosis) to ensure that this was not a confounding factor in analysis. Due to the wide variety of healthcare settings represented within the Medicaid dataset, we believe our findings will be generalizable across insured populations within the United States; however, international generalizability may be limited. This work is also unlikely to accurately represent the experiences of patients without insurance, who experience different (and generally more severe) barriers to care, especially in the case of chronic conditions^41^. Another general limitation of using Medicaid data is the amount of missing data regarding racial and ethnic identities. data was only recorded as White, Black/African American, or missing, meaning that other potential racial disparities could not be measured. Similarly, high missingness in ethnicity data rendered us unable to measure possible ethnic disparities.

In this work, we use a validated phenotype^17^ that requires laparoscopic surgery to define our endometriosis cohort. However, recent clinical guidelines recommend that treatment be explored concurrently with further diagnostic exploration if a clinical (vaginal) or imaging test (ultrasound, MRI) indicates endometriosis^5^. Therefore, some prescriptions that occur in the “pre-diagnostic” subgroup are likely meant to treat suspected endometriosis. Another limitation of this phenotype is the use of a “female” gender marker as inclusion criteria, which may exclude some transgender men with endometriosis from cohort inclusion.

The use of ATC level 3 drug classes enables us to quickly and accurately pull relevant drugs based on their pharmacological functions while remaining robust to the fact that providers in different states may vary in their precise prescription patterns^42,43^. However, ATC medication classes are intended to reflect the drug indication, which may not always be consistent with why the drug was prescribed.

### Conclusions

In this work, we document racial disparities in medication prescription practices for Black endometriosis patients; these disparities exist in several aspects of treatment and are reflective of Black patients not receiving holistic endometriosis care. We compare these disparities to a non-endometriosis population and find several drug classes where disparities are larger in the endometriosis population, indicating that these disparities are unique to chronic disease management (and potentially to endometriosis). When coupled with the fact that these prescription disparities are larger pre-diagnosis compared to post-diagnosis, it becomes important for clinicians who treat chronic, complex patients to ensure that they are communicating with their patients and prescribing medications consistently and equitably.

## Data Sharing Statement

The IBM MarketScan® Multi-state Medicaid dataset is available to license at https://www.ibm.com/watson-health/about/truven-health-analytics (see ref. 12). Code for cohort definitions is available through the OHDSI Phenotype Library and at https://github.com/elhadadlab/endochar. The underlying code for this study is available on Github at https://github.com/elhadadlab/endo_disparities.

## Supplementary Material

**Supplemental Table 1:** Difference in relative prevalence between endometriosis and comparison cohort. This table contains the difference in prevalence for the 29 ATC level 3 drug classes we analyze in our study. Associated p-values are also shared.

| **Drug Class** | **Endometriosis Cohort: Prevalence (%)** | **Comparison Cohort: Prevalence (%)** | **Difference in Prevalence (%)** | **Adjusted p-value** |
| --- | --- | --- | --- | --- |
| Other analgesics and antipyretics | 43.00 | 36.28 | 6.71 | 0 |
| Opioids | 42.09 | 29.84 | 12.24 | 0 |
| Antiinflammatory and antirheumatic products, non-steroids | 38.32 | 34.44 | 3.88 | 0 |
| Cough suppressants, excl. combinations with expectorants | 30.46 | 29.50 | 0.96 | 1.269E-89 |
| Antiemetics and antinauseants | 30.18 | 22.40 | 7.77 | 0 |
| Antidepressants | 28.33 | 22.58 | 5.74 | 0 |
| Antihistamines for systemic use | 26.62 | 32.47 | -5.84 | 0 |
| Hormonal<br>contraceptives for<br>systemic use | 26.61 | 22.59 | 4.02 | 1.863E-205 |
| Drugs for peptic ulcer<br>and gastro-oesophageal<br>reflux disease (GORD) | 22.87 | 22.06 | 0.81 | 0.0 |
| Hypnotics and<br>sedatives | 22.79 | 18.39 | 4.39 | 0.0 |
| Anxiolytics | 22.48 | 13.28 | 9.20 | 0.0 |
| Quinolone<br>antibacterials | 21.50 | 17.22 | 4.27 | 0.0 |
| Corticosteroids for<br>systemic use, plain | 19.23 | 15.14 | 4.08 | 0.0 |
| Antiepileptics | 18.64 | 16.51 | 2.12 | 0.0 |
| Progestogens | 14.79 | 11.01 | 3.77 | 0.0 |
| Drugs for constipation | 13.45 | 8.98 | 4.47 | 4.848E-134 |
| Hormones and related<br>agents | 13.42 | 13.98 | -0.56 | 8.297E-272 |
| Drugs for functional<br>gastrointestinal<br>disorders | 10.79 | 9.89 | 0.90 | 0.0 |
| Estrogens | 9.99 | 5.05 | 4.94 | 7.640E-177 |
| Antiinfectives and antiseptics, excl. combinations with corticosteroids | 8.46 | 4.49 | 3.97 | 0.0 |
| Thyroid preparations | 7.77 | 5.31 | 2.45 | 7.029E-66 |
| Urologicals | 7.28 | 5.50 | 1.77 | 2.907E-103 |
| Other systemic drugs for obstructive airway diseases | 6.33 | 5.43 | 0.89 | 0.000859 |
| Belladonna and derivatives, plain | 6.15 | 5.64 | 0.50 | 4.858E-269 |
| Iron preparations | 5.18 | 5.45 | -0.27 | 3.175E-05 |
| Other diagnostic agents | 4.25 | 4.21 | 0.041 | 2.454E-64 |
| Antifibrinolytics | 3.64 | 2.013 | 1.63 | 7.487E-50 |
| Selective calcium channel blockers with direct cardiac effects | 3.38 | 2.72 | 0.65 | 4.449E-45 |

